# Quantifying the immunological distinctiveness of emerging SARS-CoV-2 variants in the context of prior regional herd exposure

**DOI:** 10.1101/2022.03.06.22271974

**Authors:** Michiel J.M. Niesen, Karthik Murugadoss, Patrick J. Lenehan, Aron Marchler-Bauer, Jiyao Wang, Ryan Connor, J. Rodney Brister, AJ Venkatakrishnan, Venky Soundararajan

## Abstract

The COVID-19 pandemic has seen the persistent emergence of immune-evasive SARS-CoV-2 variants under the selection pressure of natural and vaccination-acquired immunity. However, it is currently challenging to quantify how immunologically distinct a new variant is compared to all the prior variants to which a population has been exposed. Here we define ‘Distinctiveness’ of SARS-CoV-2 sequences based on a proteome-wide comparison with all prior sequences from the same geographical region. We observe a correlation between Distinctiveness relative to contemporary sequences and future change in prevalence of a newly circulating lineage (Pearson r = 0.75), suggesting that the Distinctiveness of emergent SARS-CoV-2 lineages is associated with their competitive fitness. We further show that the average Distinctiveness of sequences belonging to a lineage, relative to the Distinctiveness of other sequences that occur at the same place and time (n = 944 location/time data points), is predictive of future increases in prevalence (AUC = 0.88, 0.86-0.90 95% confidence interval). By assessing the Delta variant in India versus Brazil, we show that the same lineage can have different Distinctiveness-contributing positions in different geographical regions depending on the other variants that previously circulated in those regions. Finally, we find that positions that constitute epitopes contribute disproportionately (20-fold higher than the average position) to Distinctiveness. Overall, this study suggests that real-time assessment of new SARS-CoV-2 variants in the context of prior regional herd exposure via Distinctiveness can augment genomic surveillance efforts.

## Introduction

To date, over 10 billion COVID-19 vaccine doses have been administered globally^1^, with over 200 million individuals fully vaccinated in the United States.^2^ Recent studies have confirmed that natural immunity (i.e. immunity gained through prior infection) is also highly protective and may even provide more durable protection than vaccination alone.^3–12^ Given that over 400 million COVID-19 cases have been reported worldwide (with over 78 million cases in the United States)1, it is likely that both vaccination-acquired immunity and natural immunity play important roles in the evolution of new SARS-CoV-2 variants.

Throughout the course of the COVID-19 pandemic, SARS-CoV-2 has evolved to generate new variants which harbor unique constellations of mutations (substitutions, deletions, and insertions). Some of these variants are designated as Variants of Concern (VOCs) based on evidence for increased transmissibility, increased disease severity, or reduced neutralization by vaccine-elicited sera or authorized monoclonal antibody treatments. Such variants include Alpha (B.1.1.7 and Q lineages per PANGO classification), Beta (B.1.351 and descendants), Gamma (P.1 and descendants), Delta (B.1.617.2 and AY lineages), and most recently Omicron (B.1.1.529 and BA lineages).^13^ As new SARS-CoV-2 variants evolve, it is important to estimate their likelihood of evading existing regional herd immunity and potentially transmitting highly at the community level. While the main evidence that a variant evades immune response typically relies on laboratory assays and epidemiological evidence,^14–20^ complementary approaches that provide initial evidence, as soon as new sequences are reported, could enable earlier response.

Here, we define a new metric ‘Distinctiveness’ to capture the proteome-level novelty of emerging SARS-CoV-2 sequences against all the documented regional lineages. Distinctiveness aims to quantify previous herd exposure to viral sequences that are similar to the current sequence, capturing an important factor of viral competitive fitness. This approach views viral evolution through a new lens that considers the pressure to evolve new strains harboring protein content to which communities have not previously been exposed. We show that the same lineage can have different Distinctiveness values simultaneously in different countries, as well as different Distinctiveness-contributing positions. We find that the relative Distinctiveness of emergent SARS-CoV-2 lineages is associated with their competitive fitness, as defined by the change in the lineage prevalence. We also show that epitope positions contribute disproportionately to Distinctiveness.

## Results

### ‘Distinctiveness’ as a metric to capture novelty of emerging SARS-CoV-2 sequences

Understanding the immunological novelty of a SARS-CoV-2 strain for a given population needs to take into consideration which sequences were previously seen at a regional level and for which there might exist population-level immunity. Here, we introduce a new metric ‘Distinctiveness’ of a given SARS-CoV-2 sequence based on comparison against all available sequences previously collected from the same region. Specifically, Distinctiveness is defined as the average distance at the amino-acid level between a sequence and all prior sequences (**Figure 1**; see **Methods**). Distinctiveness can be computed at the global level or at a regional level for any chosen time period. Below we compare Distinctiveness of the VOCs with contemporary sequences and investigate the relationship between Distinctiveness of a sequence and the change in its regional prevalence. For comparison, we also report the ‘Mutational load’ of the same sequences. Mutational load is simply defined as the number of mutations in the new sequence compared with the ancestral reference sequence (GenBank: MN908947.3), and as such it does not account for the entirety of SARS-CoV-2 evolution or the local prevalence of sequences.

**Figure 1.**
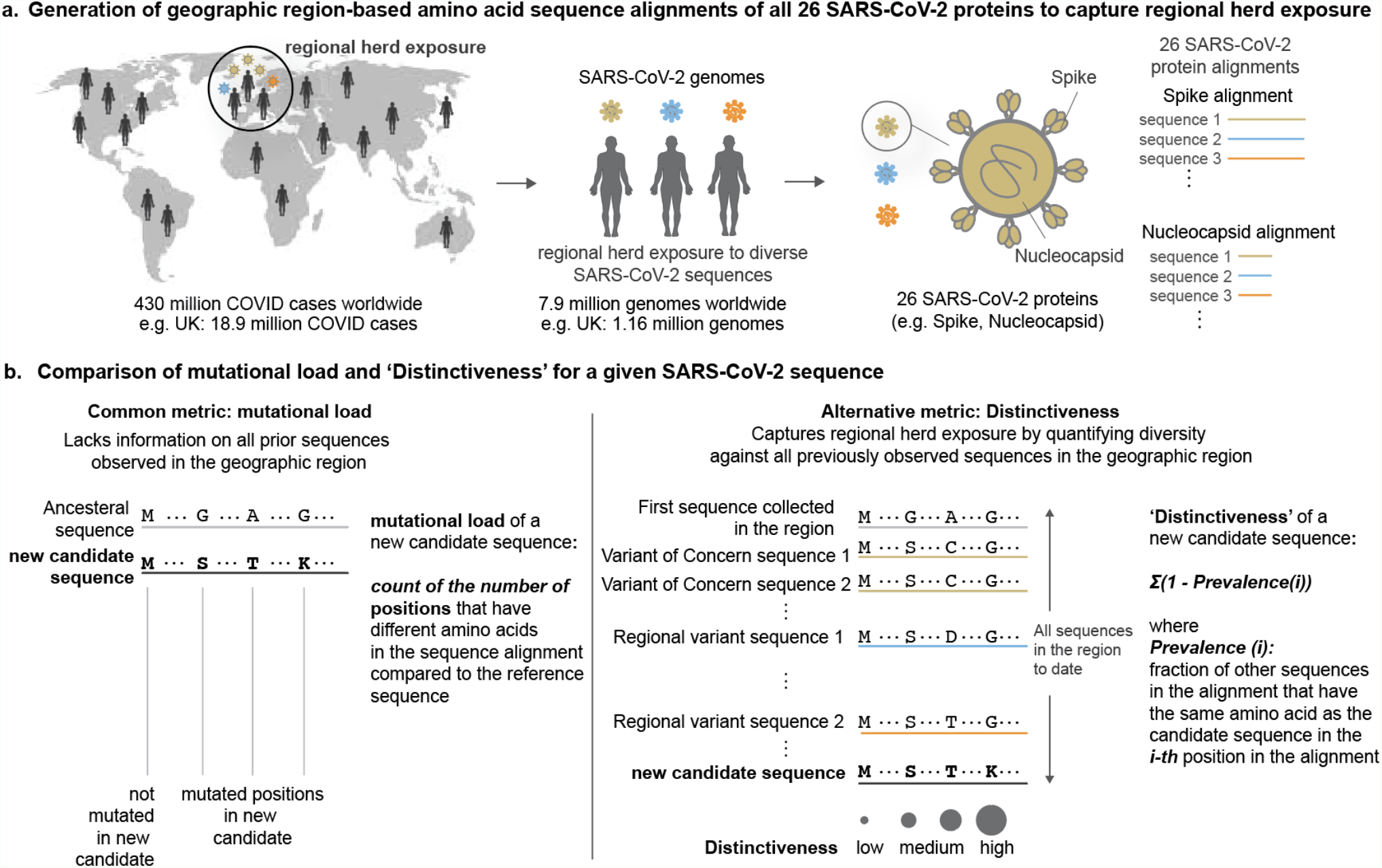
**a**. Generation of geographic region-based amino acid sequence alignments of all 26 SARS-CoV-2 proteins to capture regional herd exposure. **b**. Comparison of mutational load and ‘Distinctiveness’ for a given SARS-CoV-2 sequence.

We computed mutational load and Distinctiveness during the emergence of the VOCs in the country of their emergence. Both mutational load and Distinctiveness values of VOC sequences were significantly higher than contemporary lineages (**Figures S1,2**). For example, we consider the emergence of the Delta variant in India during January 2021. Both mutational load and Distinctiveness of the Delta variant in India were significantly higher than that of the other contemporary lineages (**Figure 2a**). This raises the question of whether Delta variant sequences were also competitive in other countries. We considered the example of Brazil, where the Gamma variant was dominant prior to the arrival of the Delta variant (**Figure 2b**). Whereas the mutational load of the Delta variant was comparable to those of contemporary lineages, the Distinctiveness of the Delta variant was significantly higher. Indeed, the Delta variant outcompeted the Gamma variant to become the dominant strain in Brazil (**Figure 2b**).

**Figure 2.**
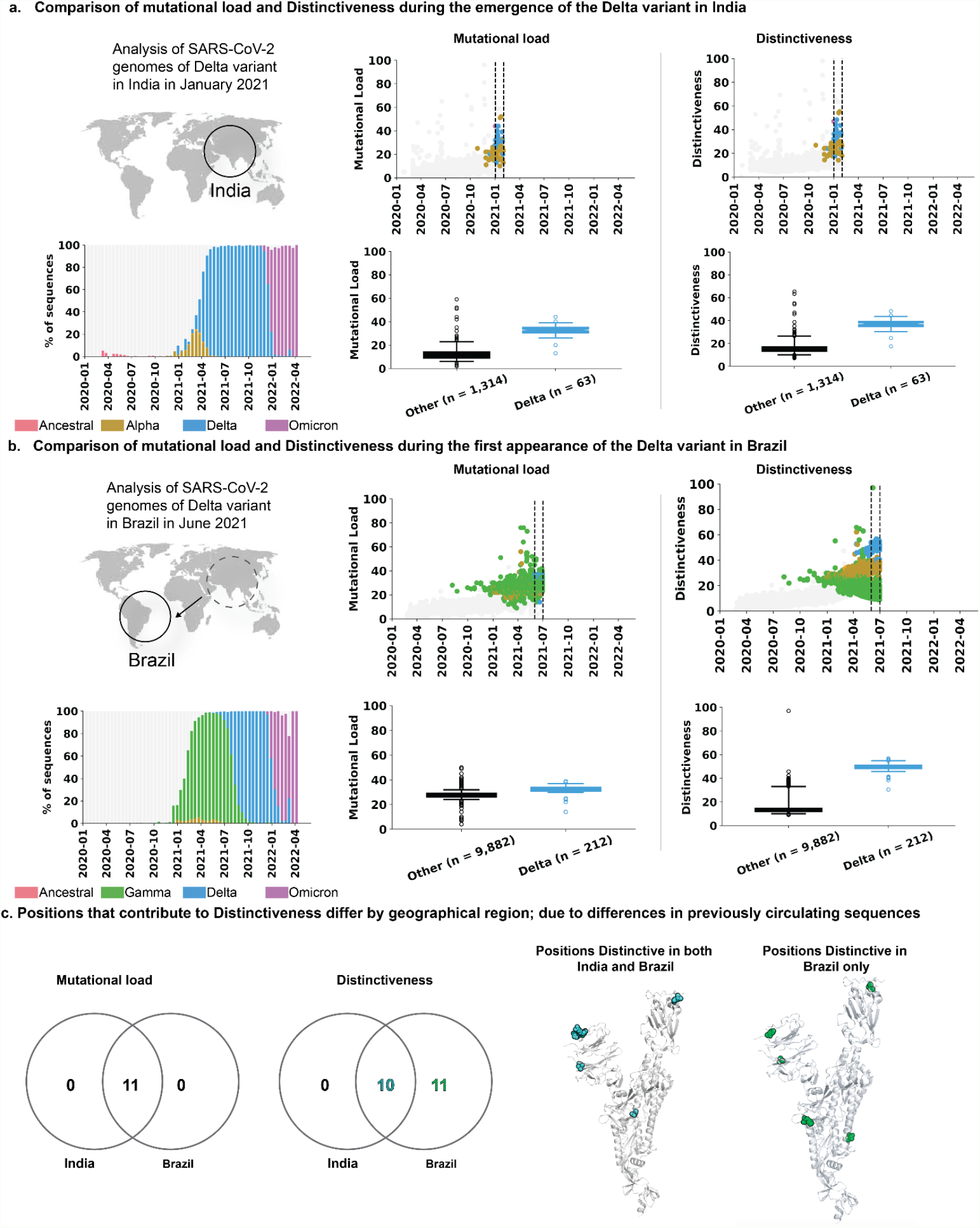
Sequence Distinctiveness as a function of time in geographical regions where VOCs first emerged. Comparison of mutational load and Distinctiveness during the emergence of the Delta variant in: **a**. India, and **b**. Brazil. **c**. Comparison of mutational load and Distinctiveness in Spike protein of Delta variant in India and Brazil. Venn diagrams compare the positions that contribute to Mutational load or high Distinctiveness (from Figure S3). The positions that have high Distinctiveness are highlighted on the protein structures of the Spike protein as spheres (PDB identifier: 6VSB). **a-b**. Sequences classified as VOCs are brightly colored dots (Alpha: brown, Beta: orange, Gamma: green, Delta: blue, Omicron: magenta) and other sequences are gray dots. Shown on the right is a comparison of the Distinctiveness values for the emerging VOC sequences and contemporary sequences, collected during the indicated time periods (inset and vertical dashed lines).

We next assessed which specific positions contribute most to the observed Distinctiveness values of the Delta variant in India and Brazil. We compared the mutational frequency and average Distinctiveness contribution for each amino acid position in the Spike protein of Delta variant sequences collected in India versus Brazil (**Figure 2c and Figure S3**). In India, where the Delta variant originated, the 11 mutated positions correspond almost exactly to the Distinctiveness-contributing positions. The only exception is the 614 position on the Spike protein. This position has not contributed to the Delta variant’s Distinctiveness as it has been highly prevalent globally (i.e. present in over 99% of SARS-CoV-2 genomes deposited in GISAID) since June 2020.^15,21,22^ Brazil, on the other hand, experienced a large wave of cases dominated by the Gamma variant before the arrival of the Delta variant. Here, in addition to the same 10 Spike protein mutations that were observed in India (**Figure S3a**), there were 11 other positions that further contributed to its regional Distinctiveness (**Figure S3b**). These additional positions correspond to known Gamma lineage-defining mutations (L18F, T20N, P26S, D138Y, R190S, K417T, E484K, N501Y, H655Y, T1027I, V1176F).

### Relative Distinctiveness of emergent SARS-CoV-2 lineages is associated with their competitive fitness

In order to examine a possible relationship between Distinctiveness and competitive fitness of SARS-CoV-2 lineages, we assessed the correlation between Distinctiveness and change in prevalence for all circulating lineages (grouped as the VOCs and a single group combining all non-VOCs) in 78 geographical regions (27 countries and 51 US states). We find that the relative Distinctiveness of emergent SARS-CoV-2 lineages is associated with their change in lineage prevalence over eight weeks (**Figure 3a, Figure S4a**) (Pearson r = 0.75). In comparison, mutational load was found to have a lower association with change in prevalence (Pearson r = 0.53). We further find that the average Distinctiveness of a lineage in a country/time window can predict future increases in prevalence (**Figure 3a**, AUC = 0.88 [0.86-0.90, 95% CI], for predicting a greater than 20 percentage point increase in local prevalence; **Figure S3b**).

**Figure 3.**
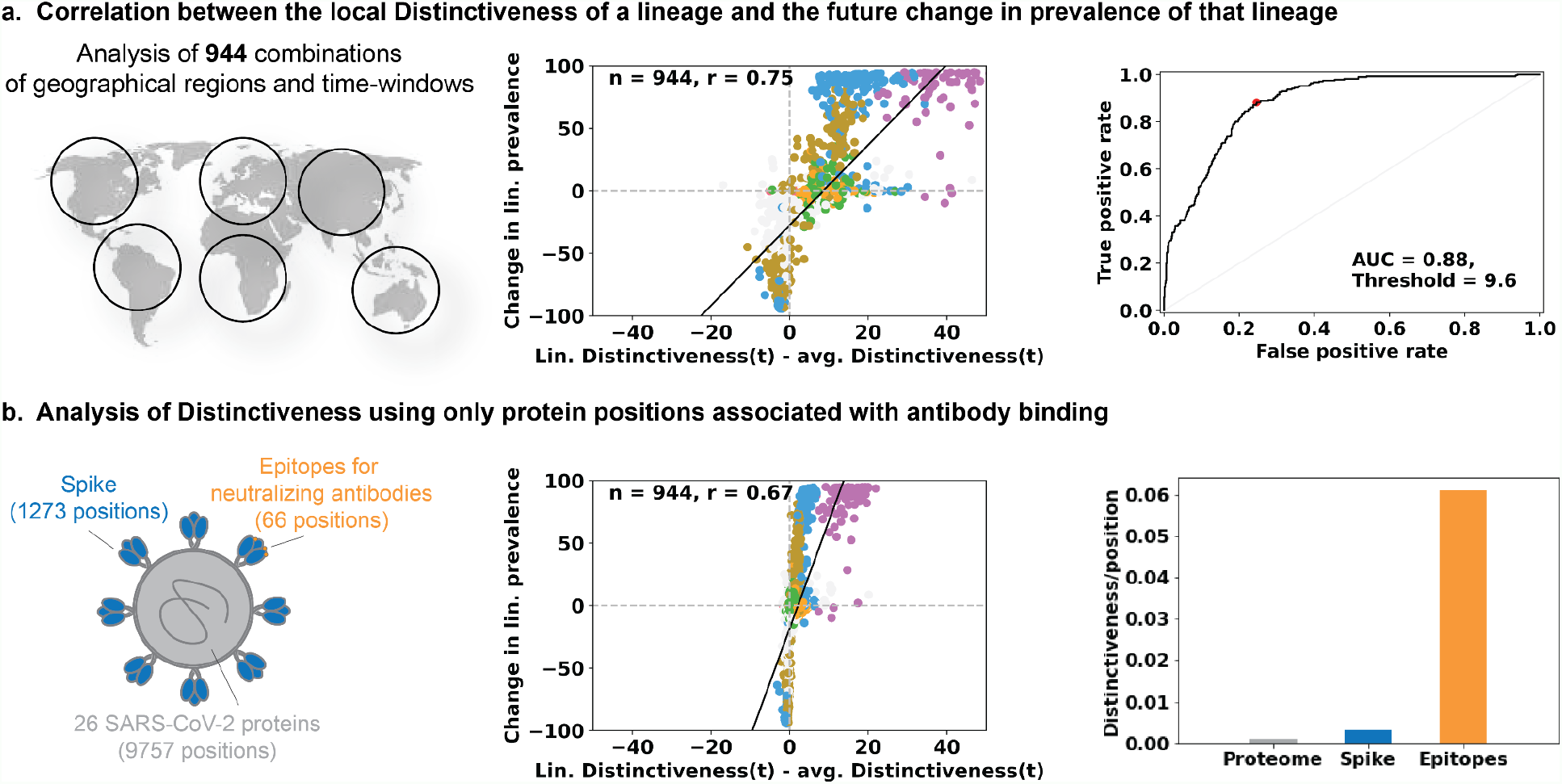
The average sequence distinctiveness as a predictor of future changes in prevalence of a lineage. **a**. Comparing the correlations of Distinctiveness of a lineage with its competitiveness in the geographic region (across 294 country/time data points). Distinctiveness of a lineage, relative to the average of all sequences that were collected from the same region during the same time, is predictive of future changes in prevalence. The ROC is shown for predicting an increase in prevalence of greater than 20 percentage points from an initial 28-day time window and a subsequent 28-day time window, starting 56 days in the future. **b**. Distinctiveness calculated for only a subset of 66 positions involved in neutralizing antibody binding (orange) retains most of the predictive capacity. These positions were found to contribute disproportionately to the overall Distinctiveness, with ∼20-fold higher average Distinctiveness as compared with average positions.

### Positions that constitute epitopes contribute disproportionately to Distinctiveness compared to the overall SARS-CoV-2 proteome

Since Distinctiveness is intended to capture the fitness of a sequence in the context of previous herd exposure to similar sequences, we next investigated Distinctiveness in the context of known immunogenic positions. Specifically, we analyzed the Distinctiveness of only Spike protein positions and for 66 epitope positions, previously associated with neutralizing antibody binding or therapeutic agent binding (**Figure 3b**). We found that Distinctiveness determined using only the 66 epitope positions was still correlated with future changes in lineage prevalence (Pearson r = 0.67). Additionally, we found that both Spike positions (average Distinctiveness of 0.007/position) and the 66 epitope positions (average Distinctiveness of 0.061/position) exhibit increased contributions to overall Distinctiveness (average Distinctiveness of 0.003/position).

## Discussion

The Distinctiveness metric defined here provides an intuitive quantification of the extent to which any viral sequence differs from other sequences that circulated previously, within the same geographical region. As such, it captures both the emergence of new amino-acid substitutions (e.g., D614G)^23^ and deletion of sequence regions that may be involved in antibody recognition,^16,24^ both of which can affect viral sequence immunogenicity and infectivity. We find that Distinctiveness can predict future changes in local prevalence of newly circulating lineages, suggesting that Distinctiveness could contribute to accurate and early identification of newly circulating lineages that are likely to outcompete other contemporary lineages. For example, analyzing diversity in the Distinctiveness at the US state-level for the Omicron variant, there are high Distinctiveness sequences in Idaho (**Figure 4**), warranting future investigation of sub-regional Distinctiveness within variants and their determinants.

**Figure 4.**
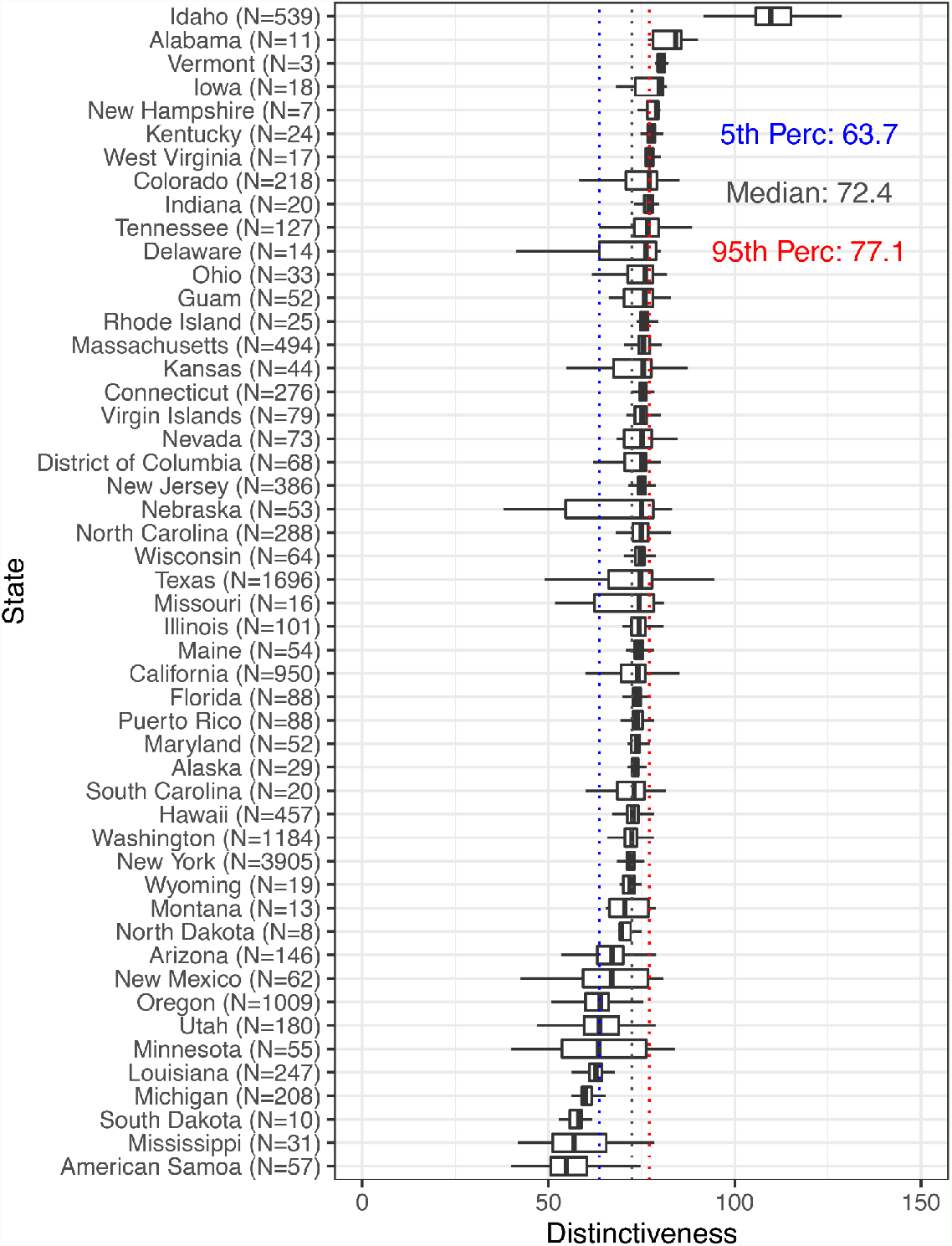
Distributions of Distinctiveness of Omicron sequences within US states after November 30, 2021.

Host immunity against SARS-CoV-2 is largely derived from two sources: vaccination and prior infection. All authorized COVID-19 vaccines utilize the Spike protein sequence from the ancestral Wuhan strain, with a slight modification (substitution of two prolines at positions 986-987) to stabilize the pre-fusion state of the protein product. These vaccines have demonstrated high effectiveness in clinical trials and various real-world studies,^17,25–40^ including against most VOCs with the notable recent exception of reduced effectiveness against the Omicron variant.^41,42^ With over 10 billion vaccine doses administered around the world, it is likely that vaccination-elicited immunity (i.e. antibody and T cell responses against the ancestral Spike protein sequence) acts as a considerable evolutionary pressure on SARS-CoV-2.^43^ The importance of natural immunity as an evolutionary pressure is highlighted by several recent studies demonstrating that prior infection confers robust and durable protection against future infection.^3–12^ Furthermore, the approach described here can be readily extended to include a correction for the durability of immunity, for example, by reduced contributions to the Distinctiveness calculation of sequences based on their collection date. We suggest that any newly emerging lineage with a combination of sequence modifications that distinguish it from the ancestral strain and VOCs that have circulated widely (or at high prevalence in a given geographic region) should be monitored closely for their potential to drive future surges.

This study has a few limitations. First, we emphasize that the Distinctiveness metric is intended as an intuitive initial evaluation of the novelty of sampled SARS-CoV-2 sequences. By design, it provides a quantification of prior herd exposure, which is a key contributor to population level immunity. However, there are additional factors, such as the functional implications of mutations and stochasticity, that determine whether a new lineage will spread widely that are not captured by our metric and that can only be captured by subsequent lab assays or epidemiological studies. Future work could combine Distinctiveness with such additional contributing factors to develop a more robust predictor of lineage competitive fitness. Second, SARS-CoV-2 genomic epidemiology is unfortunately impacted by major geographic and temporal sequencing biases. Over 55% of SARS-CoV-2 genome sequences in GISAID were isolated from infected patients in the United States or the United Kingdom, and the number of cases subjected to whole genome sequencing increased massively starting at the end of 2020. Undersampling of SARS-CoV-2 genomes in other regions and/or during earlier months of the pandemic could impact our estimations of lineage Distinctiveness. Future analysis will include SARS-CoV-2 genomes from complementary databases such as the National Center for Biotechnology Information^44^. Third, although we suggest a cut-off for Distinctiveness that can be used to monitor future emerging lineages (Figure 3), it is not clear that this cut-off will remain optimal. The future of SARS-COV-2 evolution is uncertain, and may involve smaller changes to the sequence that necessitate a lower cut-off in Distinctiveness, or a more sensitive method, such as one focussed only on key immunogenic positions. Fourth, Distinctiveness can be sensitive to sequence alignment parameters. Complementary analyses that are independent of sequence alignments are warranted to overcome this shortcoming.^45^ Finally, Distinctiveness does not take into account amino acid similarities in the sequence alignments or the recency of the SARS-CoV-2 sequences used to build the alignment. Future work should account for amino acid similarities using substitution matrices^46^ and incorporate the time of sequencing as parameters in computing the Distinctiveness scores.

In conclusion, we highlight that Distinctiveness more holistically captures the ongoing combat between viral evolution and host immunity, wherein lineages which are most distinctive from both the ancestral strain (the basis for all authorized COVID-19 vaccines) and VOCs (i.e. prior dominant strains against which natural immunity has developed) are the least likely to be neutralized by host immune responses. Distinctiveness can be considered as one important feature contributing to the competitive fitness of emerging SARS-CoV-2 variants and thus a salient factor to monitor as part of the global pandemic preparedness efforts.

## Methods

### Quantification of number of distinct positional amino acids for prevalent SARS-CoV-2 lineages

Individual substitutions, insertions and deletions for each aligned SARS-CoV-2 protein sequence along with the corresponding PANGO designation were obtained from the GISAID (https://www.gisaid.org) database, on May 3rd, 2022. We considered only sequences labeled as “complete” and “high coverage” from the GISAID data, and collected from 28 top sequencing countries (**Table S1**), this resulted in a total of 4,926,906 sequences. For the original Wuhan strain and the five VOCs (Alpha, Beta, Gamma, Delta and Omicron), the PANGO classification was obtained from the CDC website (https://www.cdc.gov/coronavirus/2019-ncov/variants/variant-classifications.html).

### Calculation of sequence Distinctiveness

For a given sequence, Distinctiveness within a geographical region of interest (i.e., a country) is defined as the average distances at the amino-acid level between that sequence and all sequences that were collected at least one calendar day before that sequence (limited by the time-resolution of the data). Specifically, for a sequence, *s*, it’s Distinctiveness, *D*(*s*), is calculated using the following formula:

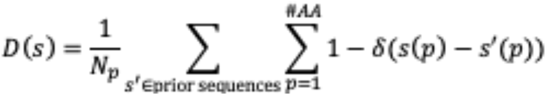

Where *N*_*p*_ is the number of prior sequences, *s’* is one specific prior sequence, the inner sum is over all pairwise aligned amino acid positions, and *δ*(*s*(*p*) - *s‵*(*p*)) evaluates to 1 if sequence *s* and *s‵* have the same amino-acid identity (one of twenty amino acids, a deletion, or a specific insertion) at position *p* and 0 otherwise. Positions of amino acids are determined relative to the Wuhan-Hu-1 reference, and insertions were treated as a single modification at the site of insertion. In cases where a nonsense mutation occurred, resulting in an early stop codon, mutations that followed this stop codon were not considered.

### Calculation of sequence mutational load

The mutational load was calculated as the number of mutations away from the ancestral Wuhan-Hu-1 sequence. Similar to in the Distinctiveness calculation, insertions were counted as a single mutation. In cases where a nonsense mutation occurred, resulting in an early stop codon, mutations that followed this stop codon were not considered.

### Calculating local prevalence of variants of concern

The local prevalence of a SARS-CoV-2 variant, as reported in **Figure 2** was calculated as the percentage of SARS-CoV-2 sequences in GISAID that were assigned to a lineage comprising that variant, during specific time windows and in specific countries.

### Correlating the Distinctiveness and changes in future prevalence of SARS-CoV-2 lineages

We correlated the average Distinctiveness of sequences in a set during a 28 day window to the change in prevalence of the corresponding set, defined as prevalence (*t*+56 to *t*+84) - prevalence (*t* to *t*+28), where *t* denotes time. For the analysis in **Figure3a** we show data points only for time periods in which one of the VOCs (Alpha, Beta, Gamma, Delta, and Omicron) first reached >5% prevalence in a given geographic region (defined as a country or US state); all variants present in the geographical region at included time windows are shown. This results in 944 data points, spanning 154 time windows in 78 geographical regions. An alternate version of this analysis, with inclusion of all available time windows (36,000 time windows spanning the same 78 geographical regions) is shown in **Figure S4** and yields similar conclusions as those described in the main text.

ROC-curves were generated from these data using Scikit-learn, using binary labels based on a minimum 20 percentage point increase in lineage prevalence for a country/time datapoint. Resulting AUC and threshold values, maximizing the sum of Sensitivity and Specificity, were found to be robust with respect to the cut-off used for labeling the data based on the percentage point increase (Figure S5). We used bootstrap resampling (10,000 samples) of the underlying data points (scatter points in Figure 3a) to estimate 95% confidence intervals on the resulting AUC values.

### Labeling of neutralizing antibody epitope sites on the Spike protein

We have abstracted antibody epitope data for Therapeutic antibodies, as tracked by NCATS (https://opendata.ncats.nih.gov/covid19/), as well as Neutralizing antibodies, typically isolated from convalescent patient sera, as encountered in the Protein Data Bank^47^. We define an antibody epitope as all residues in the antigen protein that have heavy (non-hydrogen) atoms at a distance of 4 Angstroms or less to heavy atoms of the bound antibody. When a structure of an antigen-antibody complex contains multiple instances of the interaction, such as in the case of a Spike protein trimer, and/or when several structures of the same antigen-antibody complex are available, we aggregate the binding data into a single epitope definition. We have also collected data for neutralizing antibodies as listed in Supplementary Data files provided by the Bloom and Xie labs,^48,49^ who have reported the results of single-point mutations that affect binding affinities (https://media.githubusercontent.com/media/jbloomlab/SARS2_RBD_Ab_escape_maps/main/processed_data/escape_data.csv). We have listed residues whose mutations were found to have a non-trivial effect on binding activity for a given antibody (site total escape of 0.1 or higher). These are not necessarily close in 3D structure. As structures of those antibodies with bound antigen become available, we do find good agreement, in general, and we amend the epitope definition with that derived from the 3D structure data. In a few instances, structure-derived epitopes were slightly extended based on the characterization of the epitope by the structure’s authors, and may include interactions slightly beyond the 4 Angstrom cutoff that we have employed.

Specifically, the following positions in the Spike protein were labeled as neutralizing antibody epitope sites: 13, 14, 19, 64, 66, 67, 69, 70, 75, 76, 77, 126, 140, 142, 143, 144, 145, 146, 148, 152, 153, 154, 156, 157, 158, 211, 212, 213, 243, 244, 245, 250, 251, 253, 258, 262, 346, 367, 373, 375, 376, 394, 405, 408, 410, 411, 414, 417, 440, 446, 449, 450, 452, 477, 484, 486, 489, 490, 493, 494, 496, 498, 505, 562, 1146, and 1147.

## Data Availability

All SARS-CoV-2 sequences and associated metadata were downloaded from GISAID.

https://www.gisaid.org/

## Data Availability

All SARS-CoV-2 sequences and associated metadata were downloaded from GISAID (https://www.gisaid.org/).

## Supplementary Information

**Figure S1.**
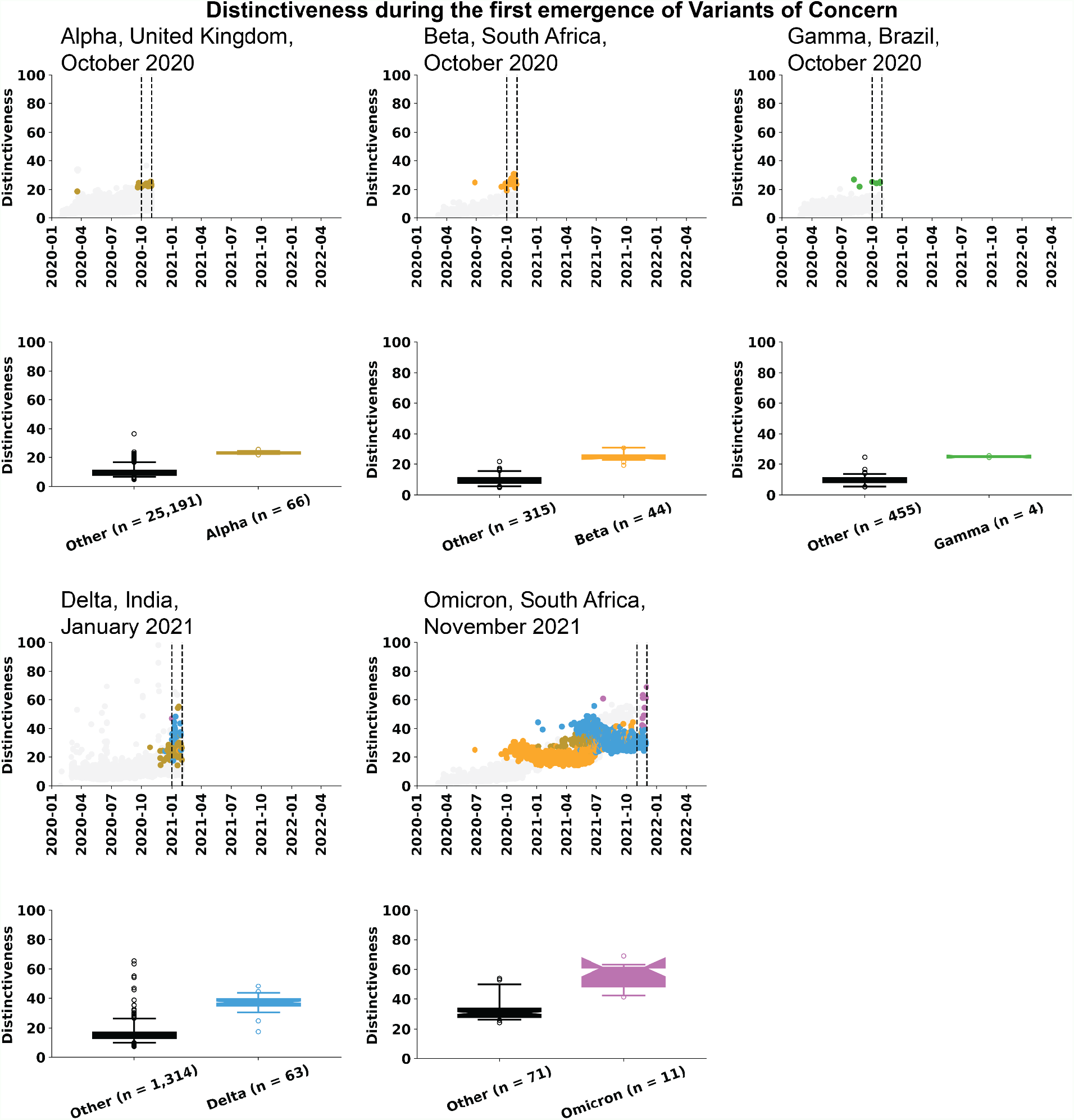
Distinctiveness of variants of concern during the time when they first appeared. In all cases, the Distinctiveness of the VOCs is significantly higher (p-value < 0.001) than that of contemporary sequences. For Alpha, Beta, and Gamma, Distinctiveness values of sequences collected during October 2020 are shown; for Delta Distinctiveness values of sequences collected during January 2021 are shown; and for Omicron Distinctiveness values of sequences collected during November 2021 are shown.

**Figure S2.**
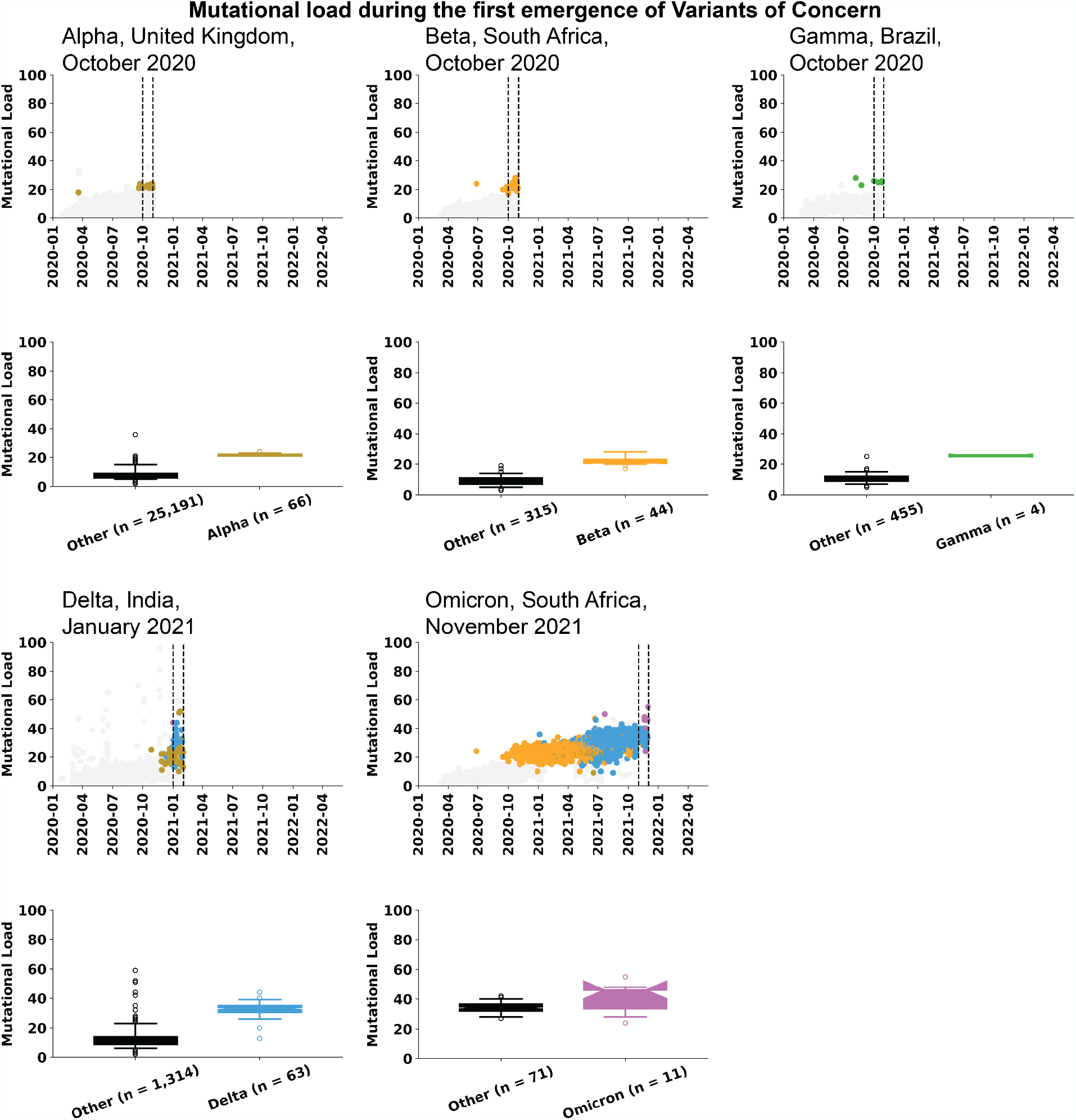
Mutational load of variants of concern during the time when they first appeared. In all cases, the Mutational load of the VOCs is significantly higher (p-value < 0.001) than that of contemporary sequences. For Alpha, Beta, and Gamma, Mutational load values of sequences collected during October 2020 are shown; for Delta Mutational load values of sequences collected during January 2021 are shown; and for Omicron Mutational load values of sequences collected during November 2021 are shown.

**Figure S3.**
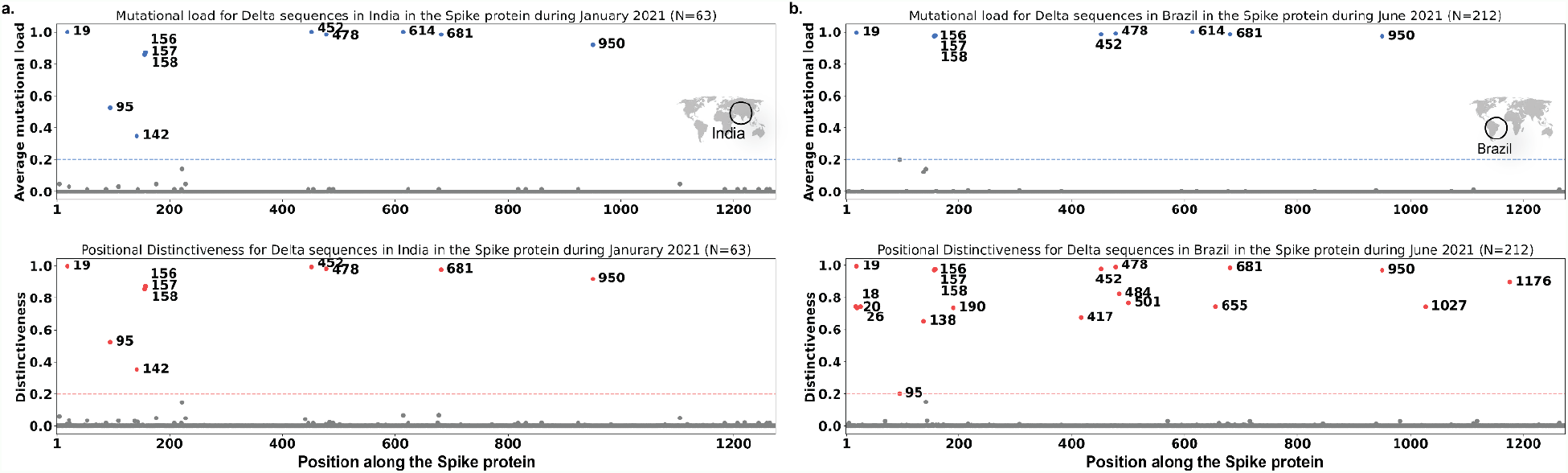
Contribution to Distinctiveness for Spike protein amino acid positions, for Delta sequences in India (**a**) and Brazil (**b**). The x-axes denote the amino acid positions in the Spike protein and the y-axes denote the average mutational load (top panel) or the Distinctiveness (bottom panel). Horizontal lines at y=0.2 denote a high threshold above which amino acid positions are labeled.

**Figure S4.**
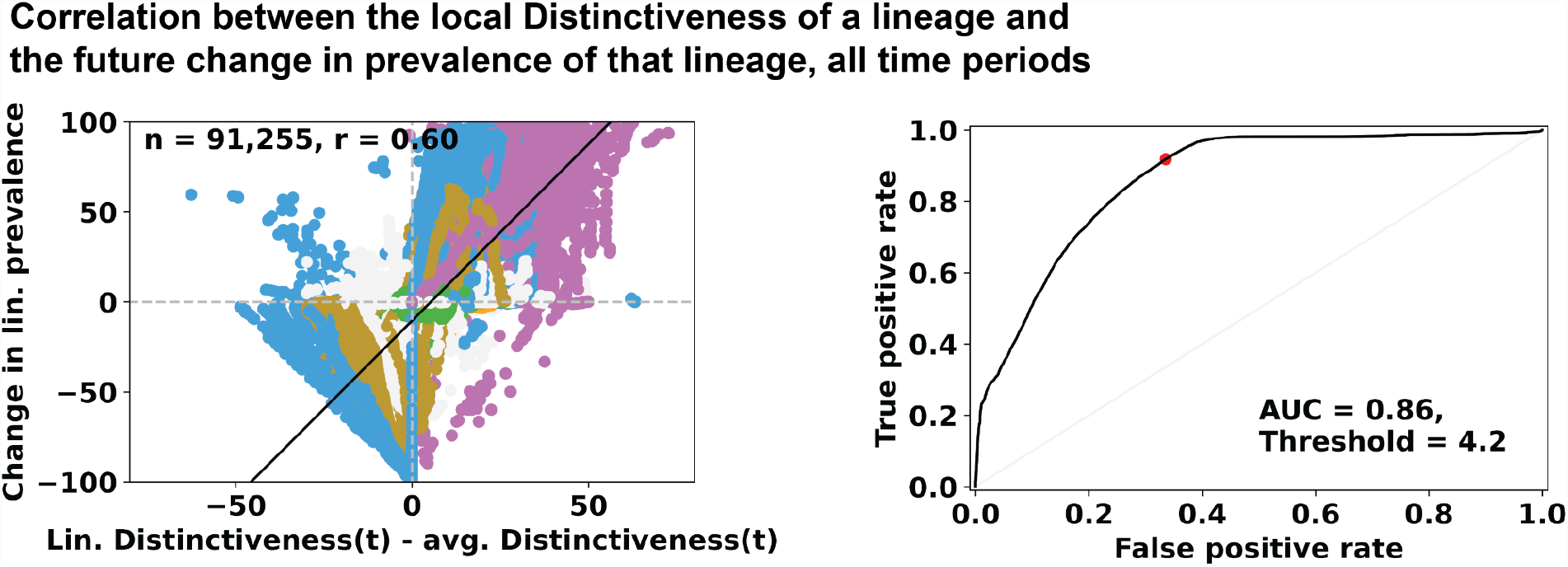
**a**. Correlation between the change in prevalence of a lineage, from the current time to 56 days in the future, without any filtering of time periods (465 time periods from 78 geographical regions). **b**. ROC for predicting an increase in prevalence of greater than 20 percentage points from an initial 28-day time window and a subsequent 28-day time window, starting 56 days in the future.

**Figure S5.**
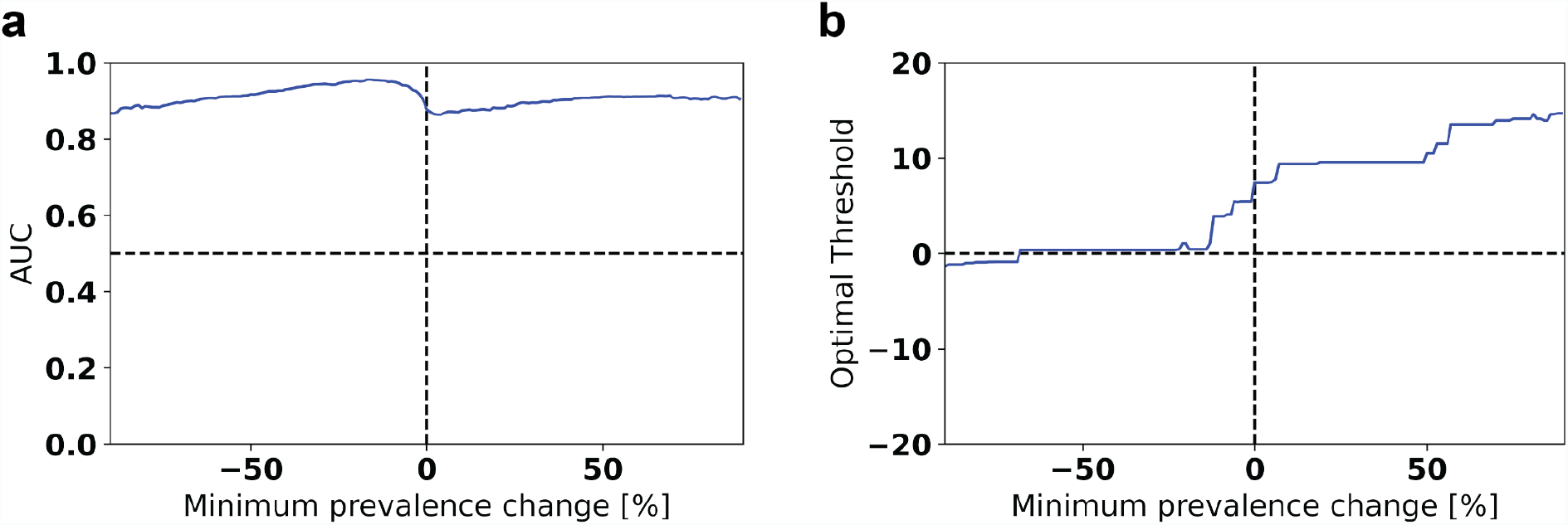
Sensitivity analysis for the prediction of future changes in prevalence from average lineage Distinctiveness. **a**. AUC for the prediction of lineages with increase in prevalence 56 days in the future from the average Distinctiveness of sequences belonging to that lineage. The minimum increase in prevalence, used to define positive labels, was varied (x-axis), and AUC values were calculated (y-axis). **b**. Optimal threshold values for a positive prediction, using average lineage Distinctiveness relative to contemporary sequences as the predictive variable, that yield the highest sum of Specificity and Sensitivity.

**Table S1.** Sequences per geographic region, included in our analysis.

| Country | Sequences from that region | US state | Sequences from that region | US state | Sequences from that region |
| --- | --- | --- | --- | --- | --- |
| United Kingdom | 1,190,215 | California | 266,055 | Louisiana | 15,156 |
| Denmark | 379,340 | Texas | 111,293 | Idaho | 14,290 |
| Germany | 290,861 | New York | 88,607 | Missouri | 13,778 |
| Japan | 182,846 | Florida | 81,862 | Kentucky | 13,775 |
| Canada | 175,571 | Colorado | 71,456 | South Carolina | 12,508 |
| France | 121,274 | Massachusetts | 63,438 | Kansas | 11,199 |
| Sweden | 116,432 | Minnesota | 59,239 | Alabama | 10,947 |
| Brazil | 84,477 | Washington | 50,713 | Iowa | 10,727 |
| Switzerland | 80,524 | Illinois | 42,502 | Wyoming | 10,454 |
| Turkey | 77,479 | North Carolina | 41,688 | Nebraska | 10,184 |
| Netherlands | 76,297 | Michigan | 41,266 | Maine | 9,355 |
| Italy | 70,548 | Utah | 40,007 | Montana | 9,069 |
| Spain | 66,644 | Pennsylvania | 36,626 | Rhode Island | 8,010 |
| India | 63,720 | Arizona | 33,575 | Hawaii | 7,889 |
| Belgium | 57,909 | Wisconsin | 32,383 | Vermont | 7,493 |
| Norway | 45,103 | New Jersey | 31,301 | Arkansas | 7,044 |
| Slovenia | 40,782 | Georgia | 30,652 | Delaware | 6,137 |
| Australia | 36,864 | Maryland | 29,086 | New Hampshire | 5,942 |
| Poland | 35,956 | Ohio | 26,979 | Mississippi | 5,781 |
| Ireland | 35,796 | Oregon | 23,820 | Alaska | 4,913 |
| Mexico | 33,546 | Tennessee | 23,401 | North Dakota | 3,384 |
| Finland | 24,028 | Indiana | 22,777 | District of Columbia | 3,192 |
| South Korea | 23,891 | Virginia | 22,199 | Oklahoma | 3,025 |
| Lithuania | 20,516 | Connecticut | 18,117 | South Dakota | 2,761 |
| Portugal | 19,349 | Nevada | 17,858 |  |  |
| Israel | 17,106 | New Mexico | 17,512 |  |  |
| South Africa | 11,362 | West Virginia | 17,045 |  |  |

